# Registered Clinical Trial Trends Evolved Differently in East Asia versus the United States during 2014 to 2023

**DOI:** 10.1101/2025.01.24.25321104

**Authors:** Eun Hye Lee, San Lee, Jae Il Shin, John P.A. Ioannidis

## Abstract

**Introduction:** East Asia, including China, Japan, South Korea, and Taiwan, has become a major hub for clinical trials. However, comprehensive comparisons between ClinicalTrials.gov and local registries remain limited. This study analyzes registered clinical trial trends in East Asia over the past decade (2014–2023) and compares them to trends in the United States.

**Methods:** We extracted clinical trial data through the International Clinical Trials Registry Platform (ICTRP), including data from local registries and ClinicalTrials.gov for China, Japan, South Korea, Taiwan, and the United States. We analyzed overall clinical trial trends and filtered randomized controlled trials (RCTs), categorizing them by location, target size, and disease category.

**Results:** China experienced rapid growth in clinical trials, surpassing both Japan and the United States in total trials and RCTs. By 2023, China led with 16,612 total trials (7,798 RCTs), while the United States registered 9,100 (4,619 RCTs). Except for China, all other countries showed some decline in recent years. Neoplastic diseases and cardiovascular and metabolic diseases were prominent in both regions, but the U.S. showed growing focus on mental health. China’s RCTs were predominantly domestic, while the U.S. maintained a higher proportion of international trials. Japan, South Korea, and Taiwan showed moderate growth, with fewer international trials than the United States. We recorded some inconsistencies and missing information across existing registries.

**Conclusion:** The study highlights the increasing prominence of East Asia, particularly China, in registered clinical trials, though with a primarily domestic focus. Improvements in existing trial registries covering East Asia are desirable.

## Introduction

East Asia, comprising countries such as China, Japan, South Korea (hereafter Korea), as well as Taiwan, is a region known for its dense population and substantial economic influence on the global stage. With a combined population exceeding 1.6 billion and one of the world’s largest and most prosperous economies, the region has become a focal point for clinical research [1,2]. Over the past decade, East Asia has ranked among the top locations worldwide in terms of studies conducted annually [3–5]. The high population density, coupled with universal healthcare systems and high-capacity hospitals, has enabled efficient recruitment and accelerated study initiation, particularly in metropolitan areas [6].

While ClinicalTrials.gov has been the primary registry used in global clinical trial analysis, the International Clinical Trials Registry Platform (ICTRP) by the World Health Organization (WHO) has also emerged as a key resource, integrating data from numerous national and regional registries [7]. In 2006, the WHO established ICTRP with the goal of providing a comprehensive view of global research to enhance transparency, improve decision-making in health care, and strengthen the validity and value of the scientific evidence base. ICTRP includes data from 18 primary registries. These registries include several focused on East Asian locations (Japan Primary Registries Network (JPRN), Chinese Clinical Trial Registry (ChiCTR), and Korea’s Clinical Research Information Service (CRiS)) [8–11]. These registries enable researchers to access a broader scope of clinical trials conducted in the region. ClinicalTrials.gov is also a data provider for ICTRP, whereas Taiwan’s local registry is not included in ICTRP. Despite the growing accessibility of multiple registries through ICTRP, comprehensive studies comparing the trial data between ClinicalTrials.gov and East Asian local registries are lacking. Doi et al. analyzed cancer clinical trials in China, India, Japan, and South Korea, focusing on registry usage and trial characteristics but limited to the cancer domain [12]. This gap in comparison restricts our ability to fully understand how clinical trial trends have evolved within East Asia, particularly in relation to the United States that has been a traditional leader in clinical research.

To address these gaps, this study aims to analyze the trends in registered clinical trials over the past 10 years in East Asia (China, Japan, Korea, Taiwan) and compare them to those performed in the United States. By integrating data from ClinicalTrials.gov and local registries in East Asia through the ICTRP, as well as separately incorporating Taiwan’s local registry [13], which is not included in the ICTRP, we classify trials based on whether they are domestic or international, their target size, and diseases addressed. We seek to provide a comprehensive overview of how the clinical trial landscape has changed in these regions compared to the United States.

## Methods

### Data extraction

To investigate clinical trial trends across East Asia (China, Japan, Korea, and Taiwan) and the United States, we extracted data from both ClinicalTrials.gov, local registries via the ICTRP and the Taiwan local registry. The ICTRP compiles clinical trial data from ClinicalTrials.gov as well as from national and regional registries [14]. The Taiwan local registry data from the Taiwan Principal Investigator Database (TPIDB), is not incorporated in the ICTRP. Instead, the data was extracted directly from the TPIDB website platform. Since the TPIDB does not provide a direct export or download function, we employed web crawling to extract the data [13]. Web crawling was conducted using the Python BeautifulSoup library (version 4.12.3) to extract individual trial information.

We retrieved clinical trial data registered over the past 10 years, between January 1, 2014 and December 31, 2023, focusing on trials conducted in China, Japan, Korea, Taiwan, and the United States. The ICTRP serves as a centralized platform providing access to trials listed on various national registries, including key East Asian registries (ChiCTR, JPRN, and CRiS). For China, Japan, and Korea, we compiled comprehensive clinical trial datasets by integrating data from both ClinicalTrials.gov and their respective local registries. In contrast, for the United States, where ClinicalTrials.gov serves as the sole national registry, trial data were exclusively extracted from ClinicalTrials.gov. For Taiwan, we integrated data by combining clinical trials registered in ClinicalTrials.gov with data extracted from TPIDB. However, TPIDB lacks trial registration dates, thus trials with reported study start dates between January 1, 2014, and December 31, 2023, were used as a proxy for registration dates. All clinical trial phases and recruitment statuses were included in the extraction.

### Data processing

Following data extraction, the clinical trial datasets from the five countries over the 10-year period (2014–2023) were processed as outlined in **Figure 1**. The ICTRP integrates data from multiple registries, which can result in some trials being registered in more than one registry. The platform automatically detects and removes some duplicates, referred to as visible duplicates. However, additional duplicates, known as hidden duplicates, may remain [15]. Therefore, after removing visible duplicates automatically, we utilized both the title and secondary ID fields to identify and remove hidden duplicates in the dataset. For trials registered in both ClinicalTrials.gov and a local registry, the earlier registration entry was manually retained [3,15]. We first determined the total number of studies and then categorized them manually based on the “study type” field to distinguish interventional studies from observational studies. Within the interventional studies, we further classified trials according to the “study design” field to identify Randomized-Controlled Trials (RCTs). Studies were categorized as RCTs if the “allocation” field indicated some form of randomization.

**Figure 1.**
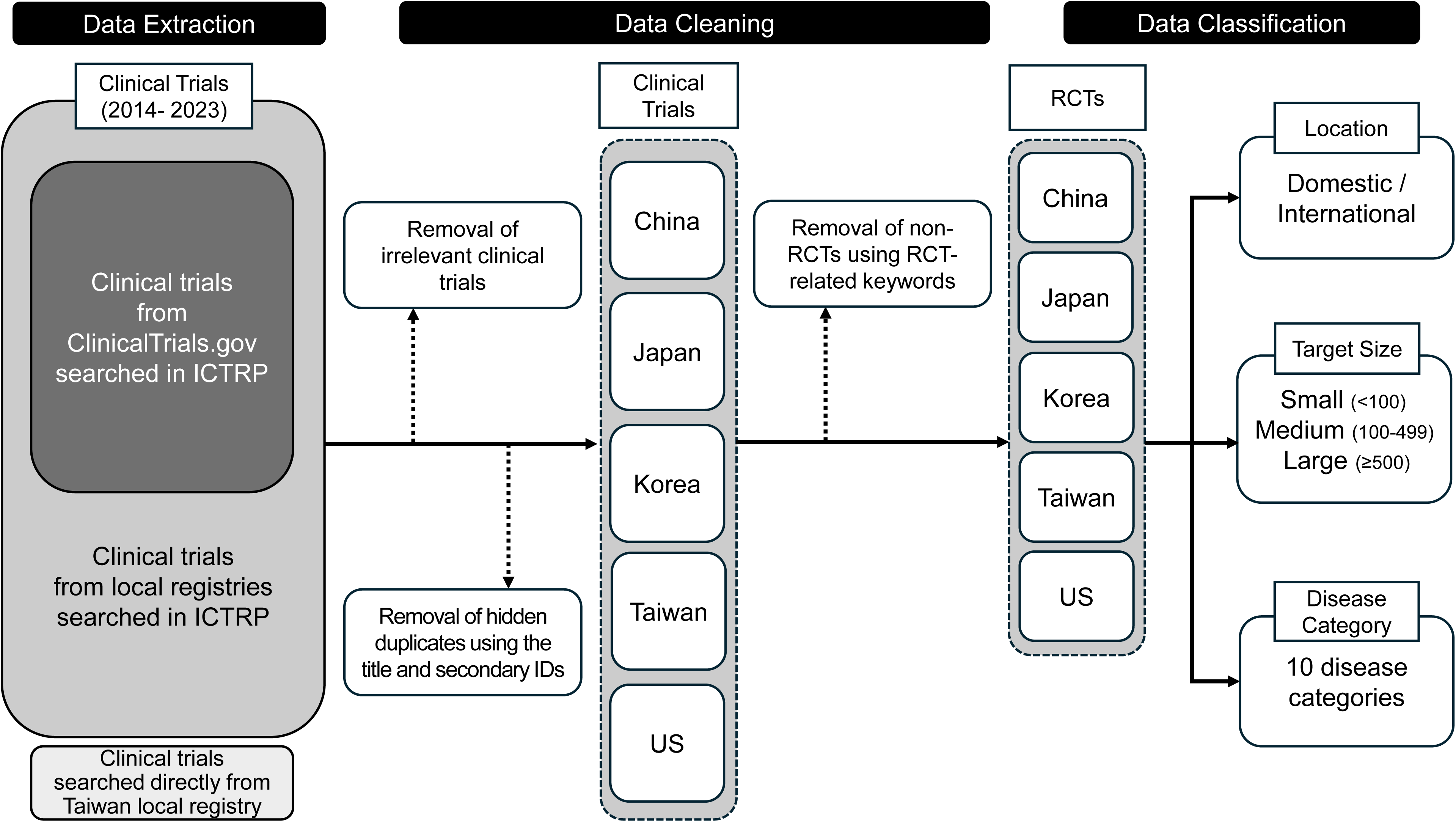
Overview of the data extraction, cleaning, and classification process of the study. ICTRP, International Clinical Trials Registry Platform; RCTs, Randomized-Controlled Trials.

RCTs were further classified manually based on location and target size. Trials conducted in only one country were categorized as “domestic,” whereas those conducted in two or more countries were classified as “international.” Additionally, target size was grouped into three categories: “small” for trials with <100 participants, “medium” for trials with 100–499 participants, and “large” for trials with 500 or more participants.

To categorize the clinical trials by disease conditions, the Medical Subject Headings (MeSH) tree structure was utilized. MeSH provide a comprehensive list of medical categories and their associated entry terms [16]. We only retained MeSH categories under the following groups: Diseases [C], Behavior and Behavior Mechanisms [F01], and Mental Disorders [F03], while other non-relevant categories were excluded [3,5]. Since MeSH categories can be highly specific, we merged related categories to create broader, more generalized disease groups. The resulting classification system consists of 9 primary disease categories, plus one category for trials not fitting into these groups (Neoplastic, Infectious, Cardiovascular and Metabolic, Nervous System, Mental, Digestive System, Respiratory Tract, Musculoskeletal, Urogenital, Other).

A language model based on Bidirectional Encoder Representations from Transformers (BERT) was employed to classify disease categories in RCTs. Specifically, PubMedBERT, a version pre-trained on biomedical literature from PubMed, was used to automate the classification of clinical trials. The 1,256 RCTs from Japan’s 2014 dataset were prioritized for initial training, with the model fine-tuned using 1,100 manually pre-classified trials. The remaining 156 trials were then classified in two separate batches to evaluate performance and re-train. For remaining trials, the model was further fine-tuned with an additional 100 manually pre-classified trials before classifying the remaining trials in a single batch. When a new set of 100 samples from Japan data was classified using PubMedBERT after training, the agreement between the manual classification and PubMedBERT classification was 95%. Classification tasks were implemented using Python (version 3.12.4, Python Software Foundation, https://www.python.org) with key libraries including pandas (version 2.2.2, https://pandas.pydata.org), PyTorch (version 2.4.1, https://pytorch.org), and Hugging Face Transformers (version 4.44.2, https://huggingface.co/transformers).

Finally, we explored the proportion of studies that were registered prospectively, the time between registration and trial start and whether prospectively registered studies had similar patterns of evolution over time as when all studies were considered.

## Results

### Trends in total clinical trials and RCTs over the last decade

As shown in **Figure 2A**, China experienced rapid growth, surpassing Japan in 2017 and the United States in 2019, with trial numbers peaking at 16,612 in 2023. In contrast, Japan’s trial numbers showed a gradual decline even before the COVID-19 period, peaking at 5,600 in 2017 and steadily decreasing thereafter. Korea and Taiwan showed more modest increases, with Korea peaking at 2,127 trials in 2021 and Taiwan reaching a high of 928 trials in the same year. Both countries experienced a slight reduction in trial numbers following the pandemic period. The United States saw steady growth, peaking at 10,595 trials in 2020 before experiencing a slight decline.

**Figure 2.**
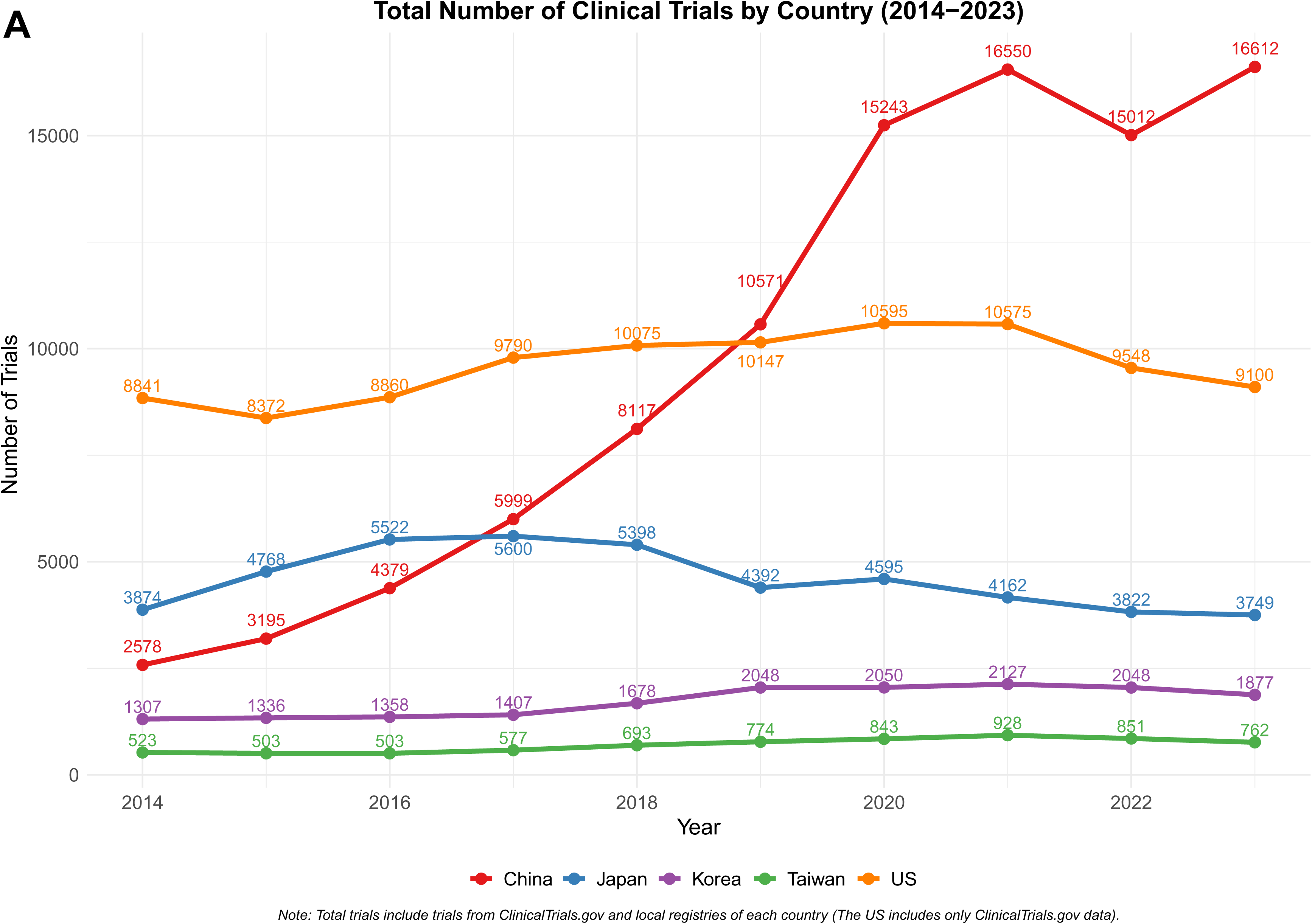
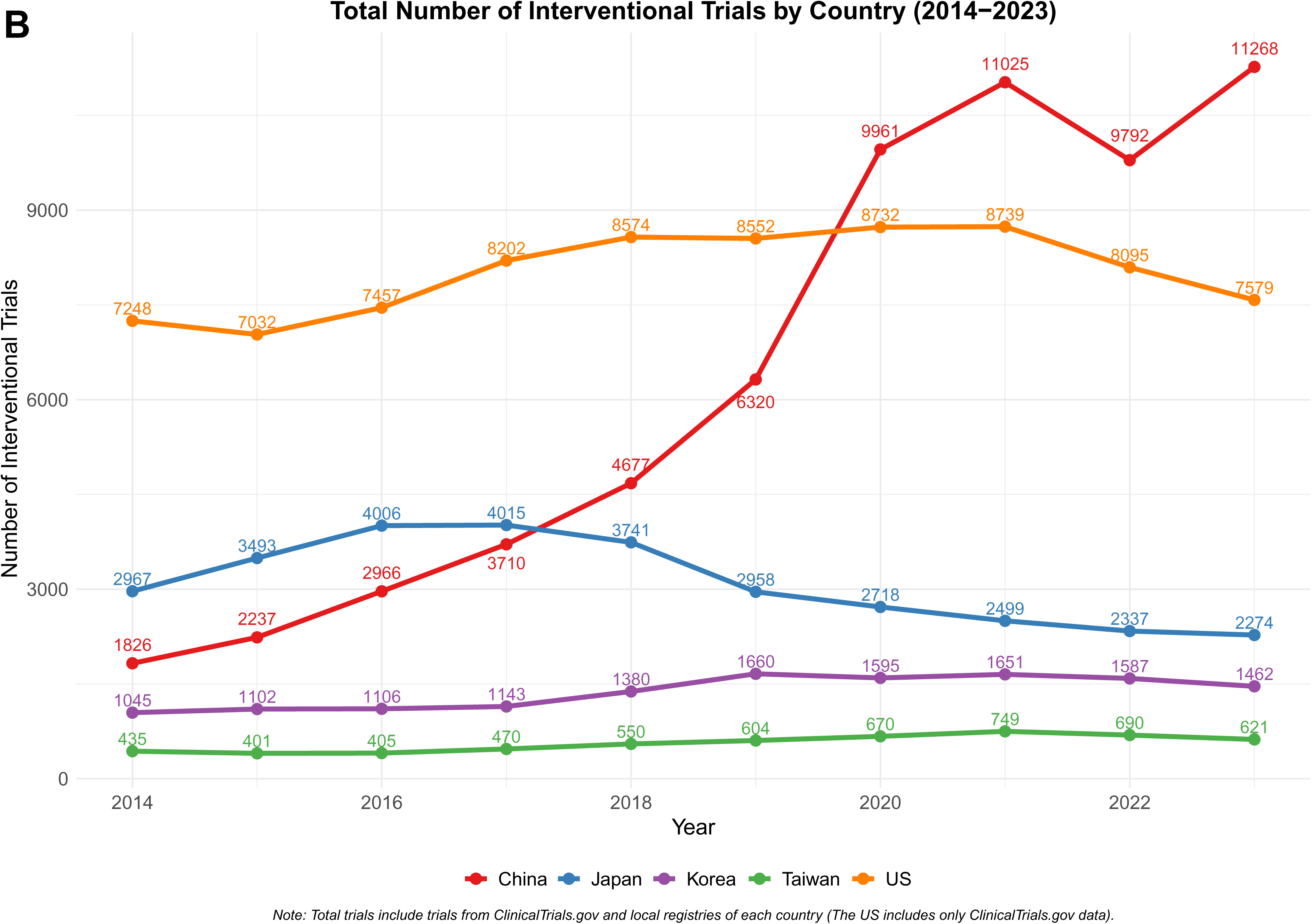
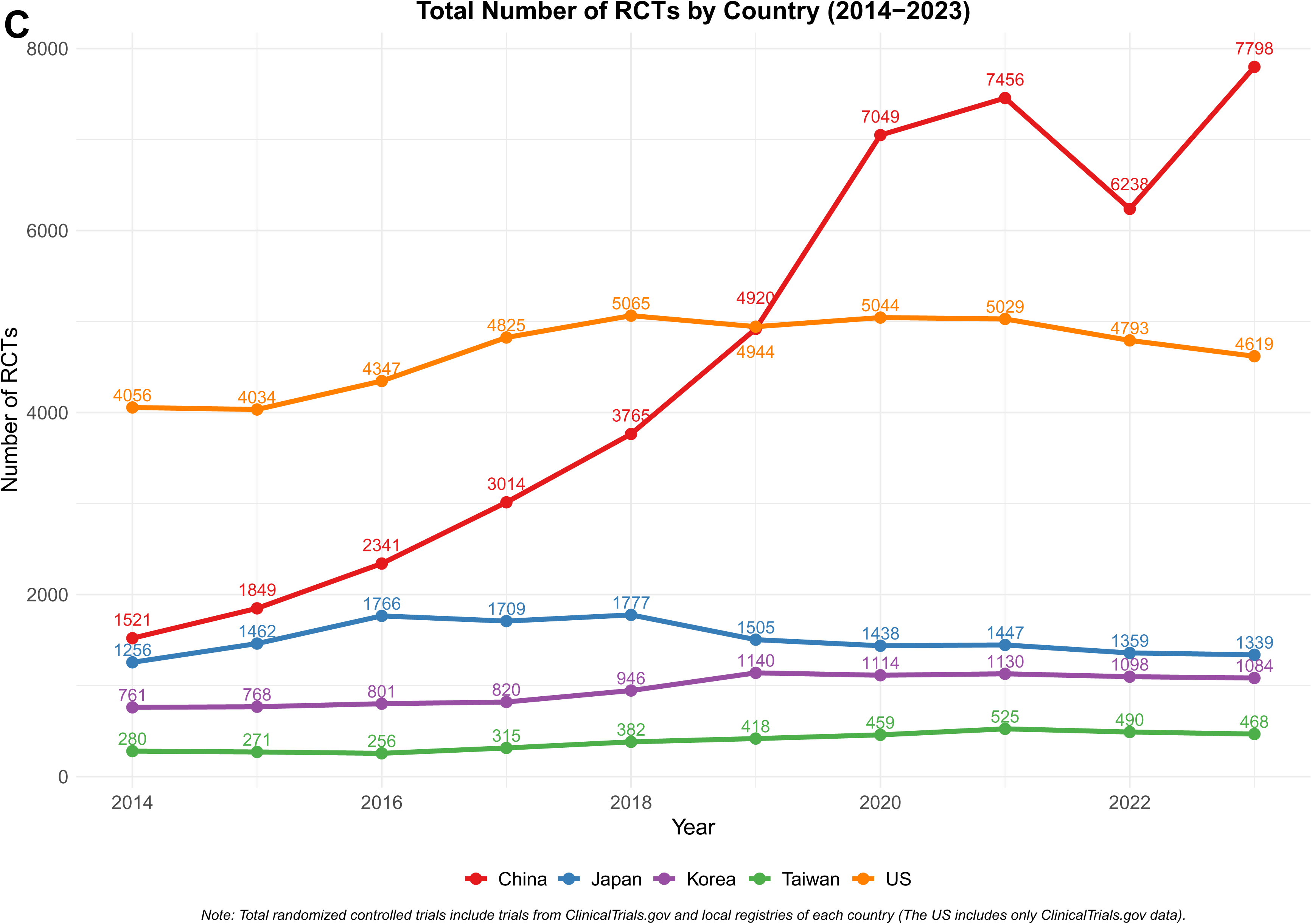
Trends in total clinical trials and randomized controlled trials in China, Japan, Korea, Taiwan, and the United States over the last decade (2014–2023). (A) Total number of clinical trials by country, (B) Total number of interventional trials by country, and (C) Total number of randomized controlled trials by country.

**Figure 2B** shows the number of interventional trials, while **Figure 2C** presents the trends in the number of RCTs. From 2014, China had more RCTs than Japan, and by 2020, it overtook the United States with 7,049 RCTs. In 2023, the number of RCTs in China peaked at 7,798, while the United States showed 4,619 RCTs during the same calendar year. Japan reached a peak of 1,777 RCTs in 2018, followed by a decline to 1,339 by 2023. Korea also showed moderate growth, peaking at 1,130 RCTs in 2021, while Taiwan’s numbers remained low, peaking at 525 in 2021 before a slight drop.

### Trends in RCT registrations by registry and location

**Figure 3** shows the distribution of registered RCTs across five countries over the last decade, categorized by registry (local or ClinicalTrials.gov) and trial location (domestic or international). In China, most trials were registered in local registries (red area), with few appearing in ClinicalTrials.gov. In 2023, there were 7,658 domestic RCTs conducted within China, with 5,841 registered on ChiCTR and 1,817 on ClinicalTrials.gov. The 140 international trials, all registered on ClinicalTrials.gov, accounted for just 1.8% of the total RCTs, reflecting the very small proportion of international RCTs. Japan followed a similar pattern, with domestic trials predominating. In 2023, domestic RCTs in JPRN accounted for 1,081, while ClinicalTrials.gov recorded only 46. International trials were more common than in China, nevertheless, with 212 (15.8%) registered in JPRN or ClinicalTrials.gov. Korea and Taiwan, likewise, registered all international RCTs on ClinicalTrials.gov. In 2023, Korea registered 1,084 RCTs, of which 185 (17.1%) were international. Taiwan registered 468 RCTs in total, with 148 (31.6%) classified as international studies. The United States, also relying solely on ClinicalTrials.gov, reported 4,619 RCTs in 2023, of which 603 (13.1%) were international.

**Figure 3.**
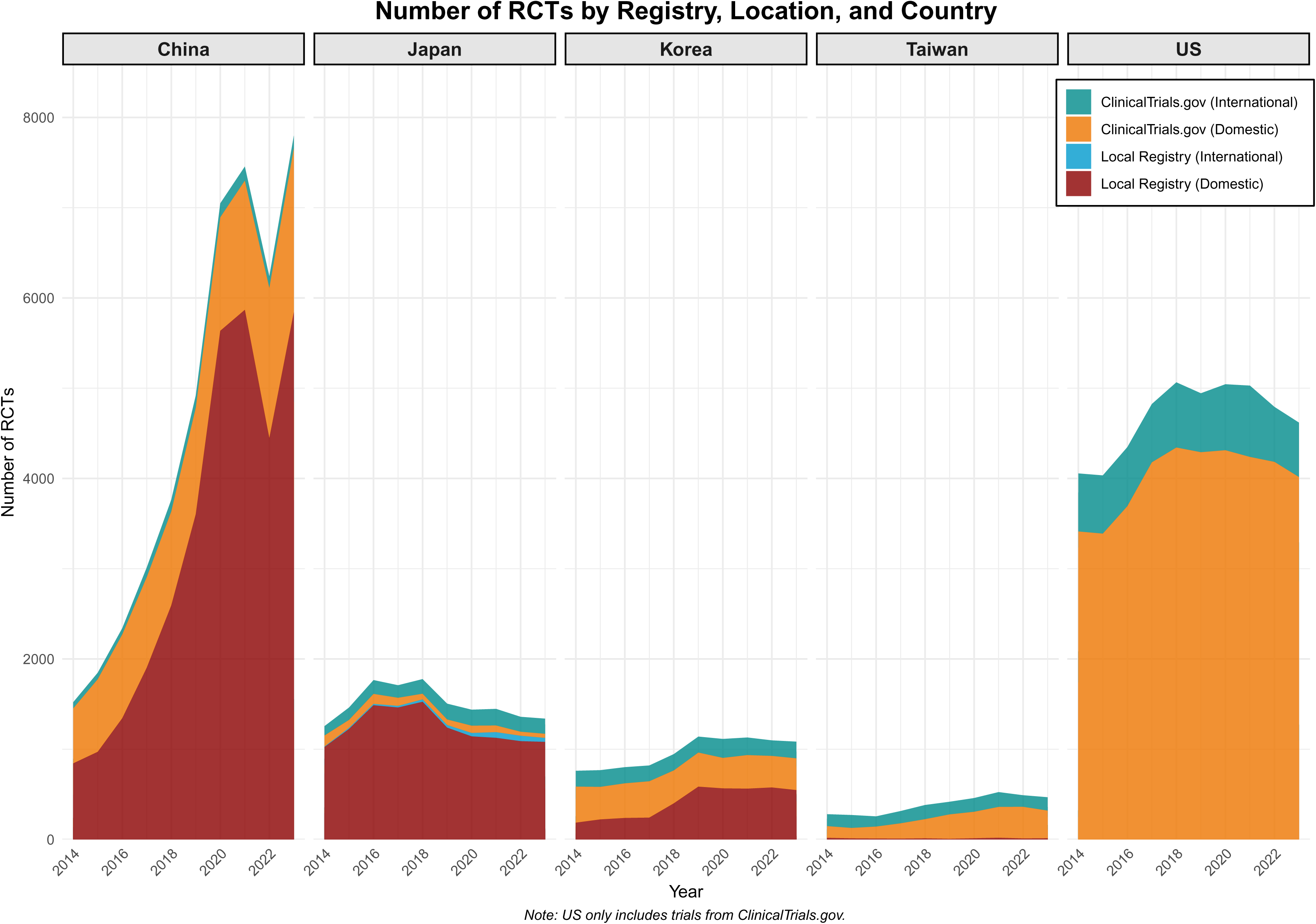
Distribution of registered randomized controlled trials by registry, location, and country over the last decade (2014–2023).

### Trends in RCT registrations by location and target size

In Japan, Korea, Taiwan, and the United States, small trials consistently formed the majority, followed by medium-sized trials, while large trials constituted the smallest proportion. In contrast, China displayed a distinct pattern, with medium-sized trials comprising the majority. Large trials accounted for 7-10% of the total in both China and the US, whereas in Japan, Korea, and Taiwan, large trials made up only 1-6%. In 2023, large trials represented 7.1% in China, 10.7% in the US, 3.2% in Japan, 6% in Korea, and 2.4% in Taiwan, reflecting a regional variation in trial size preferences (**Figure 4A**).

**Figure 4.**
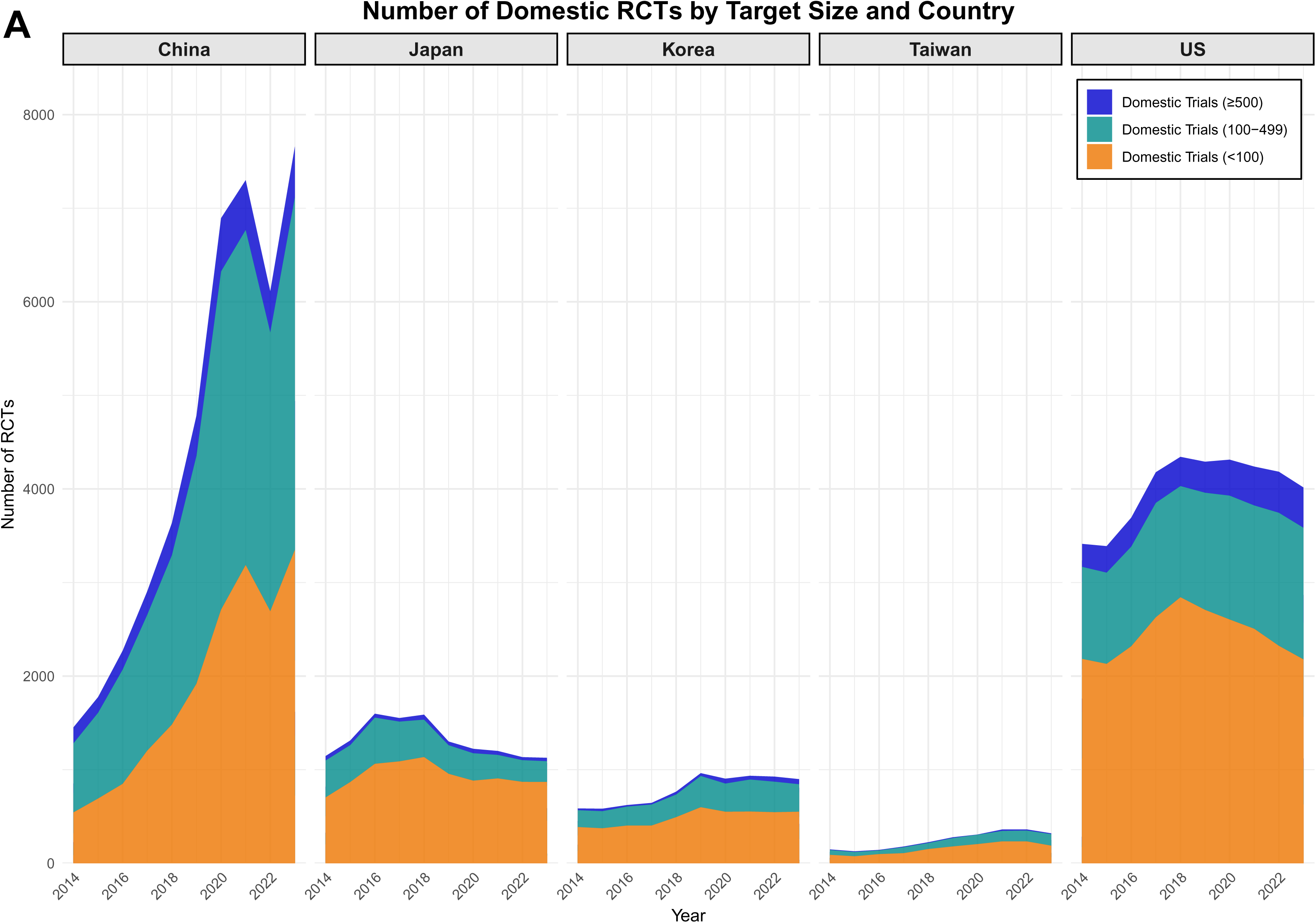
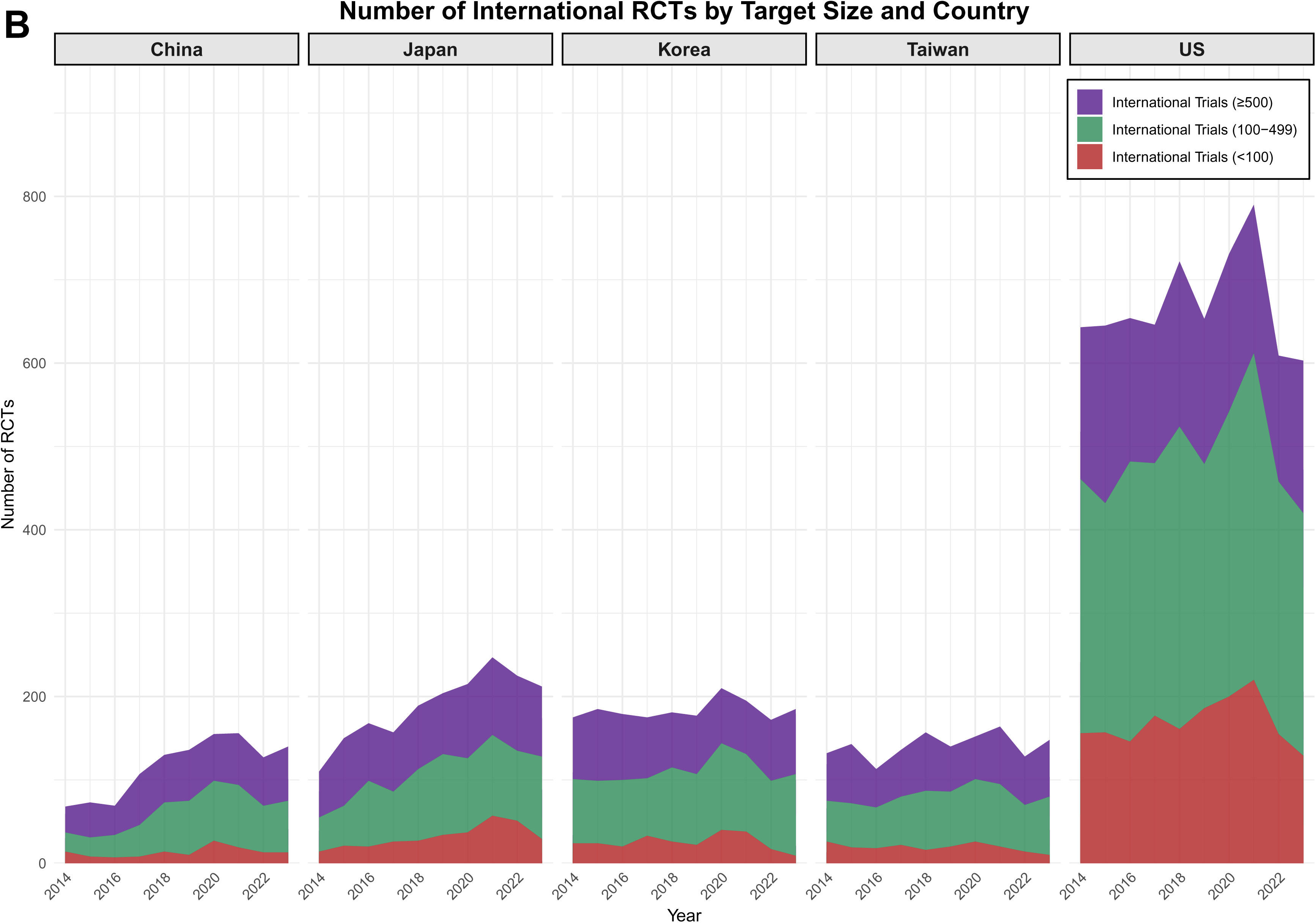
Distribution of domestic and international randomized controlled trials by target size and country over the last decade (2014–2023). (A) Number of domestic randomized controlled trials by target size and country, and (B) Number of international randomized controlled trials by target size and country.

For international RCTs (**Figure 4B**), across all five countries, medium-sized trials were the most common, followed by large-sized trials, and finally small-sized trials. However, China showed a different pattern, with large-sized trials outnumbering medium-sized trials in six out of ten years, excluding 2018 to 2021. Domestic trials dominated all countries **(Supplementary Figure 1).**

### Trends in RCT registrations by Disease Category (Figure 5)

Neoplastic Diseases and Cardiovascular and Metabolic Diseases dominate RCT registrations in all five countries. Over time, there was a noticeable increase in trials addressing mental disorders, particularly in the United States. In 2023, trials for mental disorders accounted for 16% of RCTs in the United States, making it the second most common category. Notably, the proportion of Infectious Diseases research in the United States rose from 4.9% in 2019 to 12.5% in 2020, likely driven by COVID-19-related studies, before gradually declining back to 6.5% in 2023.

**Figure 5.**
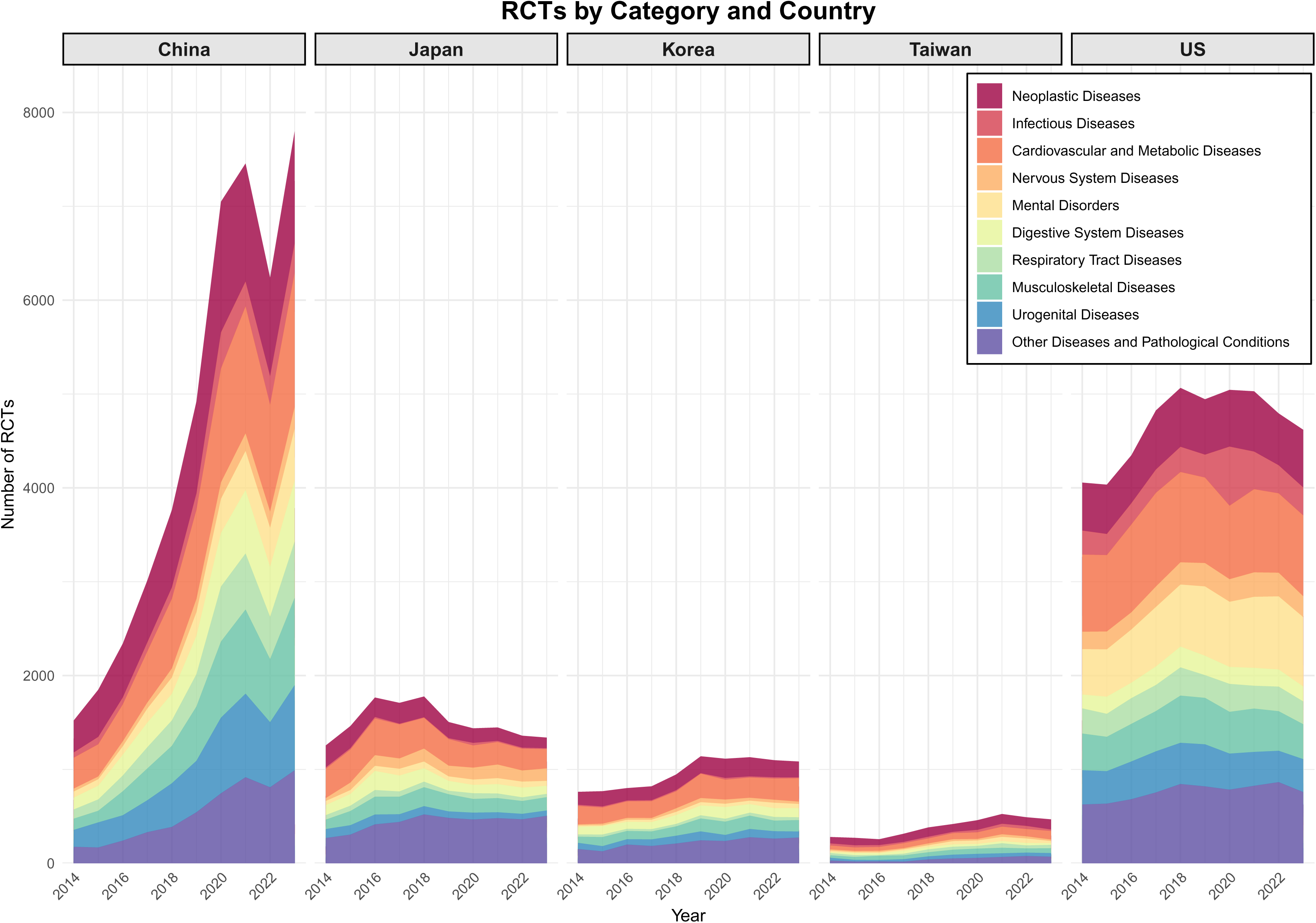
Distribution of randomized controlled trials by disease category and country over the last decade (2014–2023)

**Supplementary Figure 2** displays the proportions of RCT registrations by disease category in China and the United States, the two countries with the largest number of RCTs, for the years 2014 and 2023. In China, the share of Neoplastic Diseases and Cardiovascular and Metabolic Diseases decreased in 2023, while Nervous System Diseases and Mental Disorders increased. In the United States, Neoplastic Diseases saw a slight increase, whereas Cardiovascular and Metabolic Diseases declined, and Mental Disorders rose significantly.

**Supplementary Figure 3** shows similar data for Japan and Korea, where Neoplastic and Cardiovascular and Metabolic Diseases decreased, while Nervous System Diseases, Mental Disorders, and Musculoskeletal Diseases increased.

### Prospective registration

As shown in **Supplementary Figures 4-6**, a considerable proportion of studies were not yet registered within the first 30 days of starting and some took much longer. Prospective registration was modestly more frequent for RCTs, but many RCTs were registered well after they had started. When limited to only prospectively registered studies, the time patterns in different countries were similar to when all studies were considered.

## Discussion

Over the last decade, the clinical trial landscape in East Asia and the United States has shown distinct trends. China exhibited the most significant growth, overtaking both Japan and the United States in total registered clinical trials and RCTs. Japan, Korea, and Taiwan displayed moderate growth, although Japan saw a decline post-2017 and Korea and Taiwan have also shown small declines in more recent years. The United States experienced steady growth in clinical trials, with peaks during the COVID-19 pandemic, but a slight decline afterward. The increase in China’s trial numbers including RCTs reflects the country’s growing significance in clinical research.

The rise in China’s clinical trials and RCTs was largely driven by domestic trials registered in ChiCTR. By 2023, China had far outpaced the United States in the number of domestically conducted trials, yet international trials in China remained rare, accounting for <2% of total registrations. Conversely, the United States maintained a higher proportion of international RCTs, (13.1% of its RCTs in 2023). The concentration of Chinese trials within local registries suggests an inward focus, while the United States continues to maintain a more globally collaborative research environment.

Target sample size and disease category analysis revealed distinct patterns between regions. In China, medium-sized trials dominated both domestic and international trials, while large trials were more prevalent in international settings. The United States, by contrast, showed a more even distribution across all trial sizes. However, it is likely that many trials in all countries remain unregistered, with estimates ranging from as low as 10% to as high as 60% [17–21] and smaller trials, in particular, may have higher proportions of unregistered studies [18]. If so, this missingness may affect also the shape of the observed sample size distributions. One may speculate then that small trials may be at higher risk of remaining unregistered in China. If so, the number of trials from China may be substantially larger than observed. The growth in registered trials may reflect not just a growth in the number of trials conducted but also in the likelihood that a conducted trial is registered.

Neoplastic Diseases and Cardiovascular and Metabolic Diseases were prominent topics in both China and the United States, reflecting global priorities in addressing high-burden conditions. The United States appeared to experience a surge in clinical trial activity during the COVID-19 pandemic, likely driven by therapeutic and vaccine research needs. Following this peak, the slight decline in trial numbers may reflect the difficulties imposed by pandemic conditions in running clinical trials [22,23]. Currently, there is also a shift back toward pre-pandemic research priorities with infectious disease trials decreasing close to pre-pandemic levels [24]. Notably, by 2023, there was an increasing focus on mental disorders, likely reflecting the growing awareness of post-pandemic mental health challenges. Conversely, China has only recently started to expand its research into mental and nervous system disorders, indicating that while the country is broadening its research efforts, there may still be a lag in addressing mental health conditions.

Many studies, including even RCTs, were not registered prospectively. Authors may register these studies late, when they realize that registration may enhance their chances of publication in many journals. However, the value of such retrospective registration may be questionable.

A recent analysis of clinical trial trends in BRICS nations (Brazil, Russia, India, China, South Africa) [25], highlighted the growing importance of local registries via the WHO’s ICTRP platform. The study noted a narrowing gap in trial activity between BRICS and G7 nations, emphasizing the increasing role of emerging economies like China and India in drug development. However, this study included all clinical trials from each country registered in ICTRP without distinguishing between those registered in ClinicalTrials.gov and other local registries. This did not allow to compare the characteristics and patterns of trials across different registries. Our study contains further detail by classifying trials based on registry, location, and target sample size and includes a comparison with the United States. This allows for a more comprehensive understanding of China’s evolving role in global clinical trials. Other comparative studies [5,12], have examined differences between ClinicalTrials.gov and non-ClinicalTrials.gov databases, but these were limited by certain disease such as depression or cancer research focus.

A critical issue that emerged during our analysis was the need for better quality control in trial registries. Problems such as missing data, inconsistent formatting, and data entry errors were prevalent, across local registries compared to ClinicalTrials.gov [26,27]. One of the significant issues was the difficulty in determining study completion status. When data was extracted from various registries into the ICTRP platform, differences in registry formats led to the inability to retrieve study completion status, with only “recruitment status” information being available. When accessing the original registry URLs allowed for direct review of each registry, it was easy to find study completion information on ClinicalTrials.gov. However, for other registries, it was challenging to identify clear study completion status or dates. Thus, we were not able to comparatively assess result reporting rates for the registered trials in different countries. Even when study completion is reported in some registries, it is unknown whether the information is accurate and complete. Similarly, previous comparative studies between the European Union Clinical Trials Register and ClinicalTrials.gov also highlighted inaccuracies in completion data [28]. Additionally, inconsistent formatting across registries, including variations in how information was recorded, posed significant challenges in data standardization for cross-country comparisons. E.g., categorizations related to study design, such as allocation and randomization, varied greatly between registries. In ChiCTR, target sample sizes for the control group and multiple intervention arms are presented in text format, not provided as a total number. This required additional effort during data extraction. Without proper standardization of integrated trial data and stronger quality controls, the risk of inaccurate or incomplete trial registrations increases, potentially leading to misinterpretations. Therefore, enhanced registry protocols and appropriate quality controls are crucial, not only to ensure data accuracy and completeness but also to foster global collaboration and transparency in clinical trials.

This study has several limitations. First, removal of hidden duplicates using title and secondary IDs may not have been entirely sufficient, potentially leaving some duplicates in the dataset. Second, the target sample size classification may not accurately reflect the actual sample size due to discrepancies in trial reporting. Third, our disease classification may have oversimplified the research focus, limiting our ability to capture more detailed trends. Furthermore, probably many trials still remain unregistered and this may even vary per country, sample size, disease condition, and type of intervention.

In conclusion, standardized and integrated clinical trial platforms are needed across regions. As East Asia, particularly China, continues to expand its trials agenda, future efforts should prioritize improving data standardization in trial registration and strengthening registry quality control. This may facilitate data integration, verification, analysis, and international collaboration.

## Ethics statement

The study was not subject to institutional review board approval because it exclusively utilized open-source trial information from ClinicalTrials.gov and local registries via the ICTRP and TPIDB, without including any individual participant data.

## CRediT authorship contribution statement

**Eun Hye Lee:** Conceptualization, Methodology, Data Curation, Formal Analysis, Writing – Original Draft, Visualization

**San Lee:** Conceptualization, Methodology, Data Curation, Formal Analysis, Writing – Original Draft, Visualization

**Jae Il Shin:** Supervision, Writing – Review & Editing

**John P.A. Ioannidis:** Conceptualization, Methodology, Funding Acquisition, Project Administration, Supervision, Writing – Review & Editing.

## Data availability

The data supporting this study are available in an open-access repository at https://osf.io/f58y7/.

