## Supplementary materials for "Registered Clinical Trial Trends Evolved Differently in East Asia versus the United States during 2014 to 2023"

**Supplementary Figure 1.** Combined Distribution of domestic and international randomized controlled trials by target size and country over the last decade (2014–2023).

**Supplementary Figure 2.** Comparison of randomized controlled trials by disease category in China and the US in 2014 and 2023.

**Supplementary Figure 3.** Comparison of randomized controlled trials by disease category in Japan, Korea, and Taiwan in 2014 and 2023.

**Supplementary Figure 4.** Trends in prospective trial registration rates by country over the last decade (2014–2023). **(A)** For all clinical trials, and **(B)** for all RCTs.

**Supplementary Figure 5.** Trends in number of the prospective trial registration by country over the last decade (2014–2023). **(A)** For all clinical trials, and **(B)** for all RCTs.

**Supplementary Figure 6.** Distribution of time differences between trial registration and first participant enrollment, integrating data over the last decade (2014–2023). **(A)** For all clinical trials, and **(B)** for all RCTs.

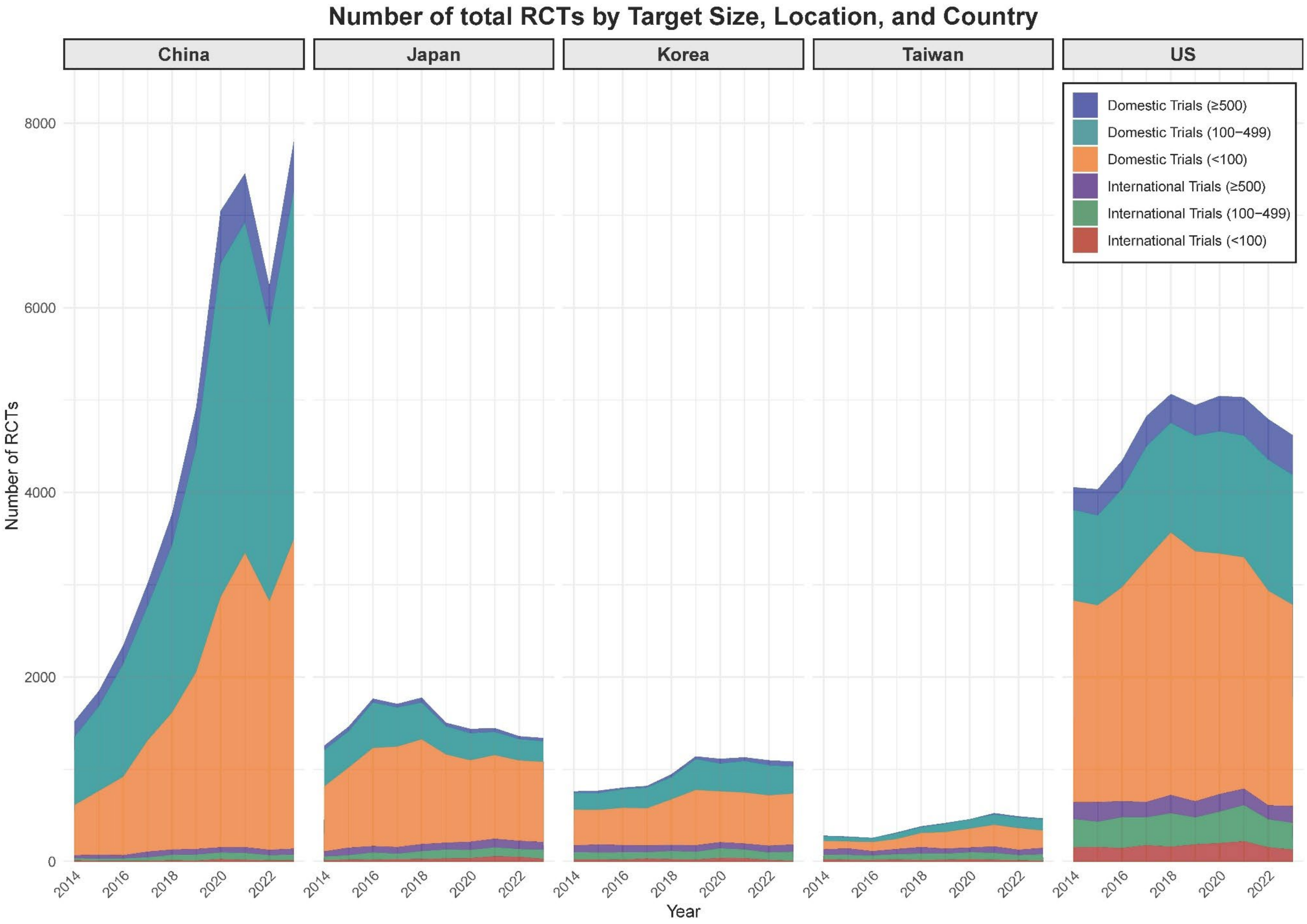


**Supplementary Figure 1.** Combined Distribution of domestic and international randomized controlled trials by target size and country over the last decade (2014–2023).

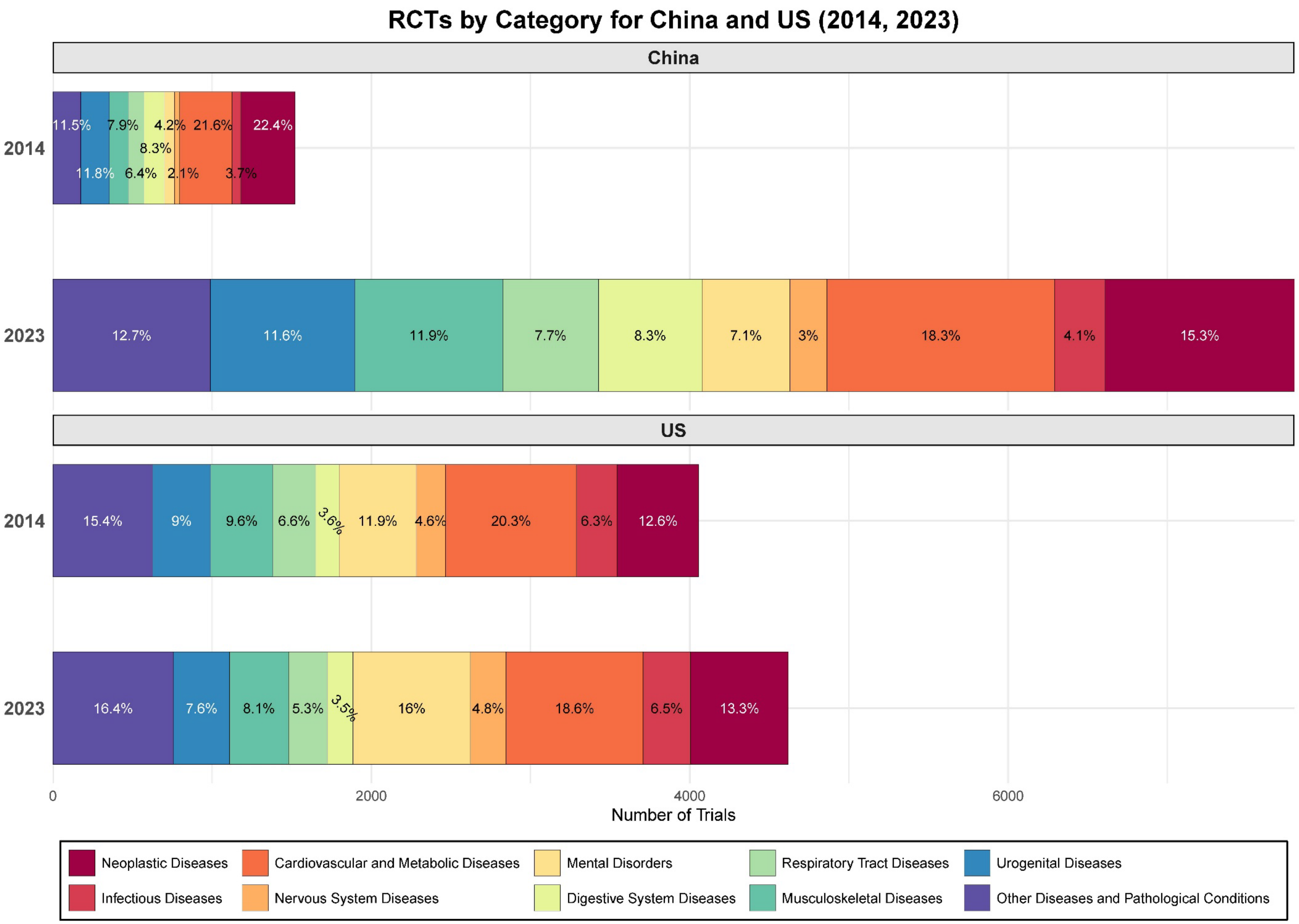


**Supplementary Figure 2.** Comparison of randomized controlled trials by disease category in China and the US in 2014 and 2023.

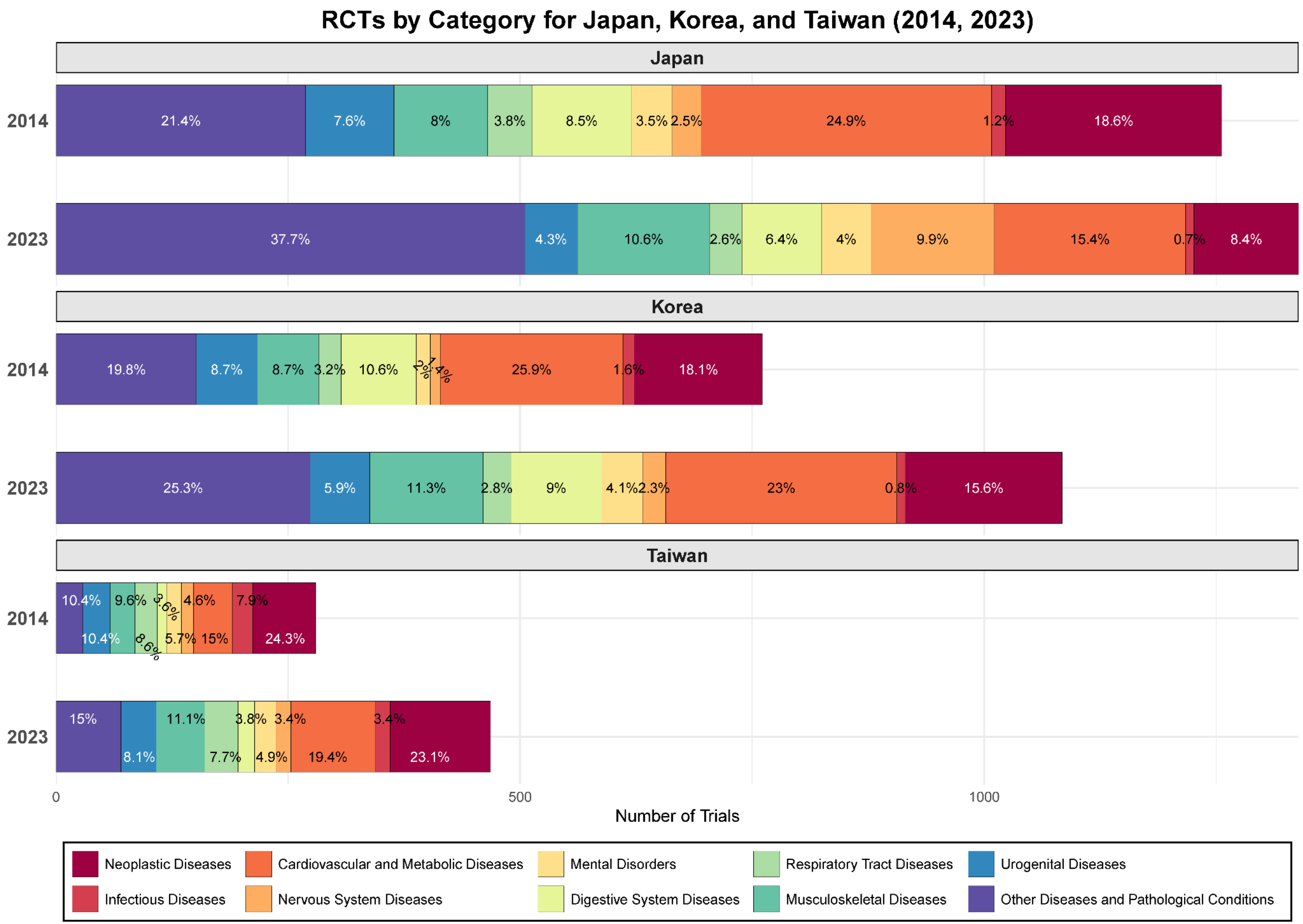


**Supplementary Figure 3.** Comparison of randomized controlled trials by disease category in Japan, Korea, and Taiwan in 2014 and 2023.

### *Trends in Prospective Registration of All Clinical Trials*

The trends in prospective registration among all clinical trials, including RCTs, were examined. Prospective registration was defined as registration occurring within 1 month of the trial start date to account for incomplete start dates and potential processing delays in the registry [1-3]. The proportion of prospective registration was analyzed by country and year, and for each country, the overall trend of the time difference between trial registration and first enrollment over ten years was visualized. As the Taiwan local registry, TPIDB, does not provide information on this time difference, TPIDB data was excluded from the prospective registration analysis for Taiwan.

As shown in **Supplementary Figure 4(A),** prospective registration proportions for all clinical trials increased in the United States from 71% in 2014 to 82.5% in 2023, with a plateau in recent years. China showed a rise from 66.8% in 2014, peaking at 85.4% in 2020, before falling to 77.8% in 2023. Japan remained stable around 70%, while Korea and Taiwan exhibited lower rates, with initial increases followed by declines. **Supplementary Figure 4(B)** highlights that prospective registration proportions were generally higher for RCTs, despite more pronounced fluctuations in Taiwan.

**Supplementary Figure 5** illustrates trends in the number of prospective trial registrations by country from 2014 to 2023, showing data for all clinical trials as well as RCTs. Compared to Figure 2(A) and (C), which show the total number of all clinical trials and RCTs, the trends in prospective registration trial numbers in Supplementary Figure 5 exhibit similar patterns.

The time differences between trial registration and the first participant enrollment are detailed using data integrated over the last decade in **Supplementary Figure 6.** A notable proportion of trials being retrospectively registered can be attributed to differences in national clinical trial registration regulations, as well as sponsor or funder requirements [4]. Additionally, many journals now require trial registration numbers as a prerequisite for submission, leading to retrospective registration of studies that were initially unregistered until their results were ready for publication [5].

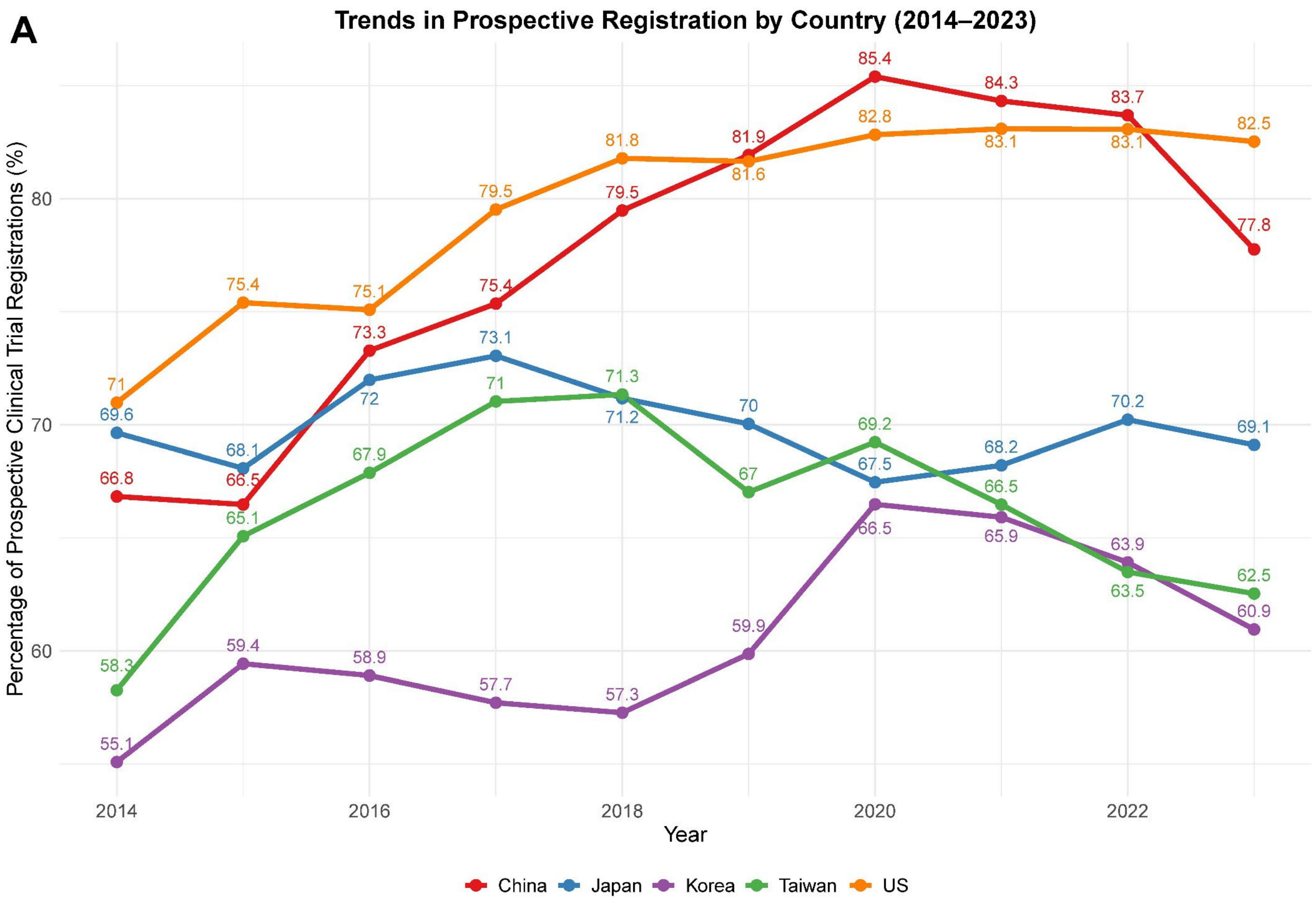


*Note: Taiwan data are only based on ICTRP data where date of registration and date of enrollment are available.*

**Supplementary Figure 4.** (A) Trends in prospective trial registration rates by country over the last decade (2014–2023) for all clinical trials.

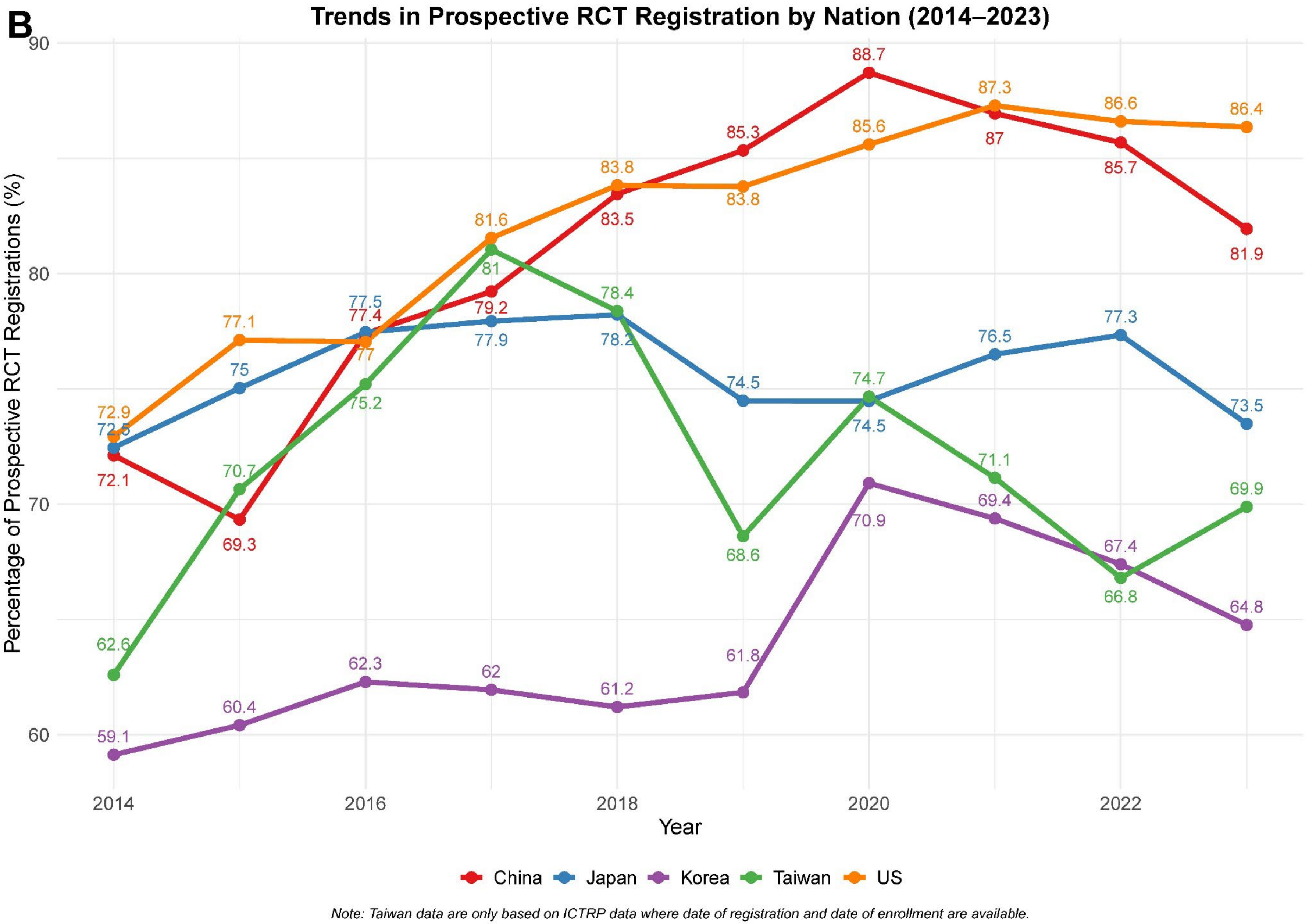


**Supplementary Figure 4.** (B) Trends in prospective trial registration rates by country over the last decade (2014–2023) for all RCTs. RCT, Randomized Controlled Trial.

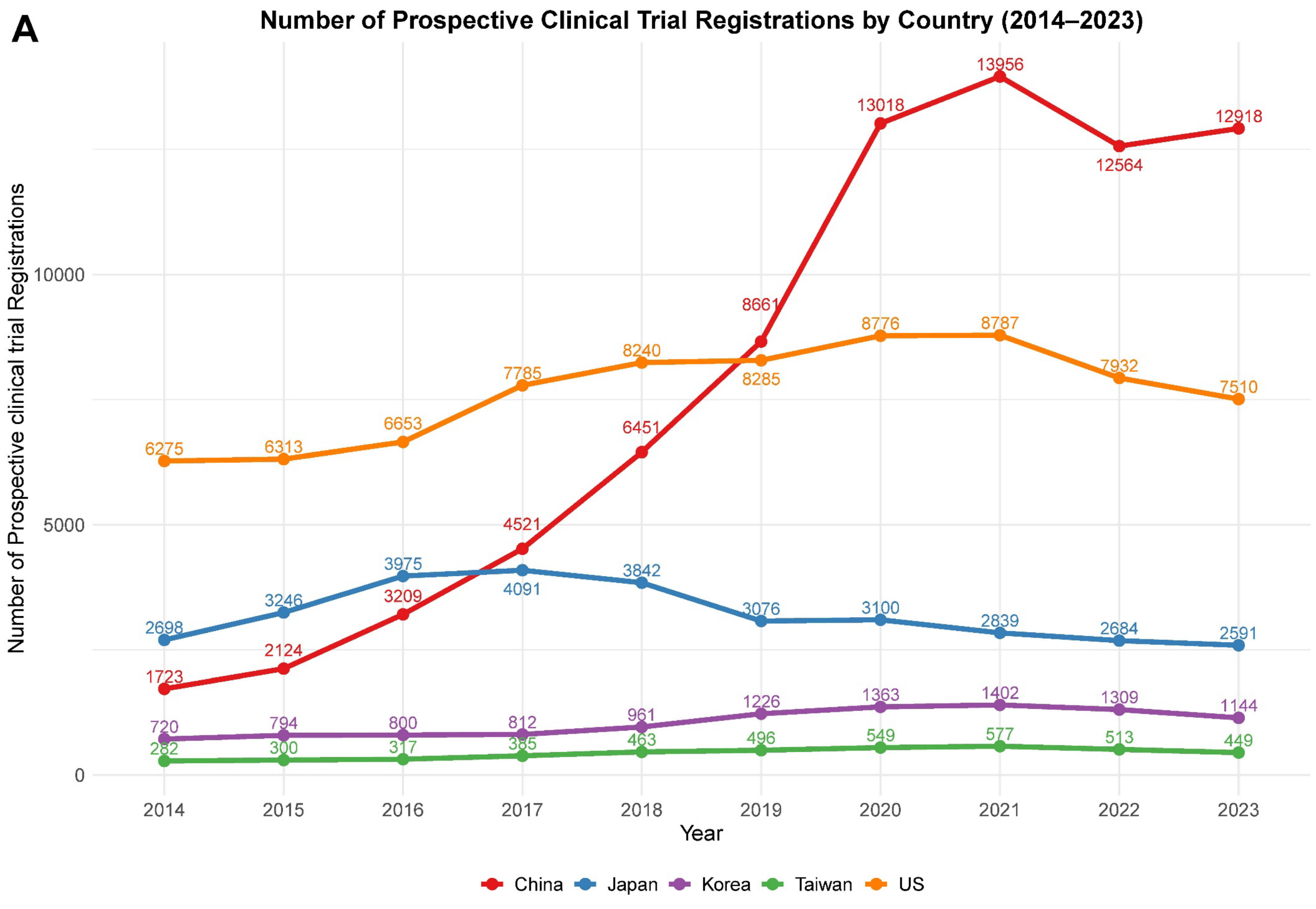


*Note: Taiwan data are only based on ICTRP data where date of registration and date of enrollment are available.*

**Supplementary Figure 5.** (A) Trends in number of the prospective trial registration by country over the last decade (2014–2023) for all clinical trials.

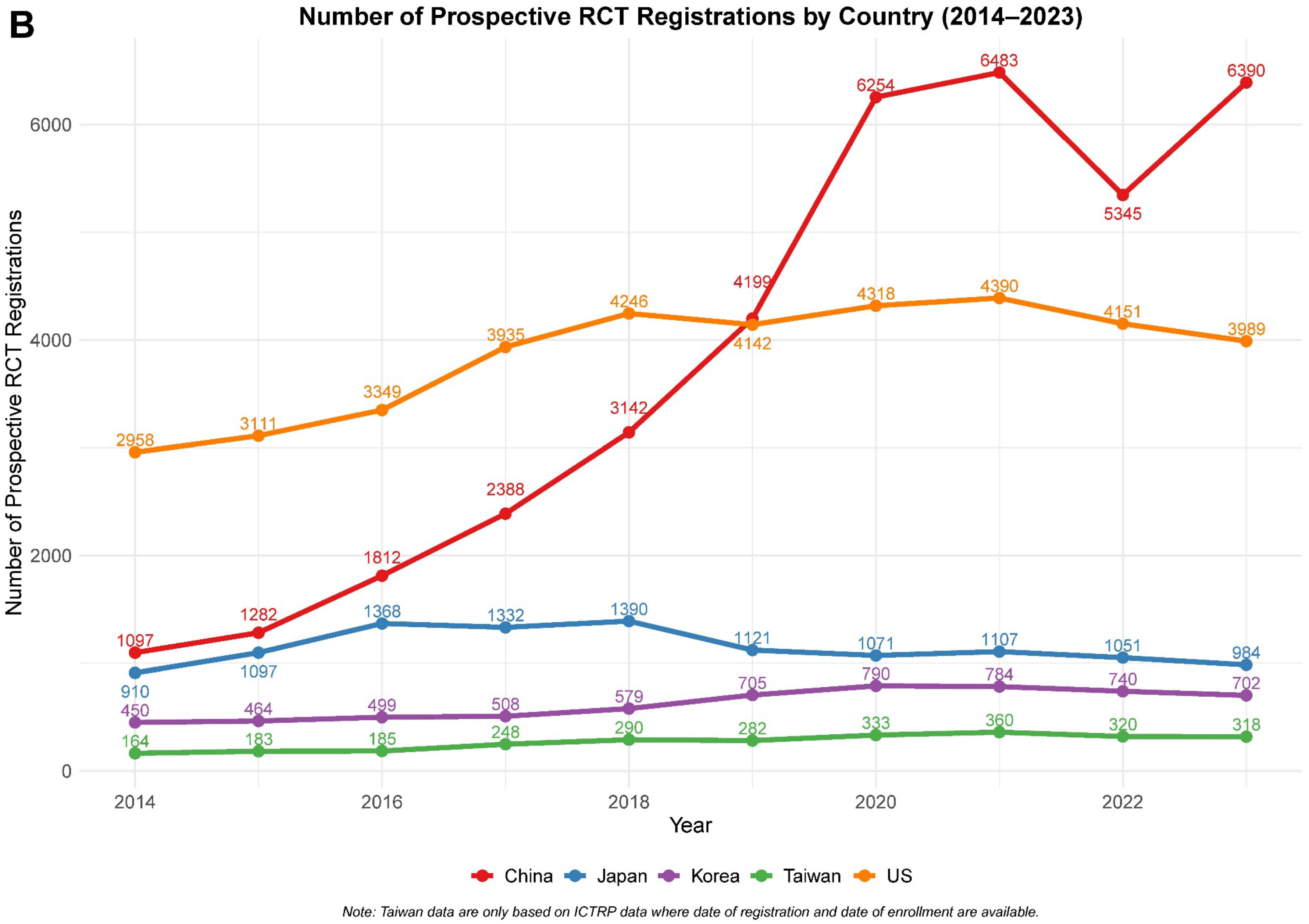


**Supplementary Figure 5.** (B) Trends in number of the prospective trial registration by country over the last decade (2014–2023) for all RCTs. RCT, Randomized Controlled Trial.

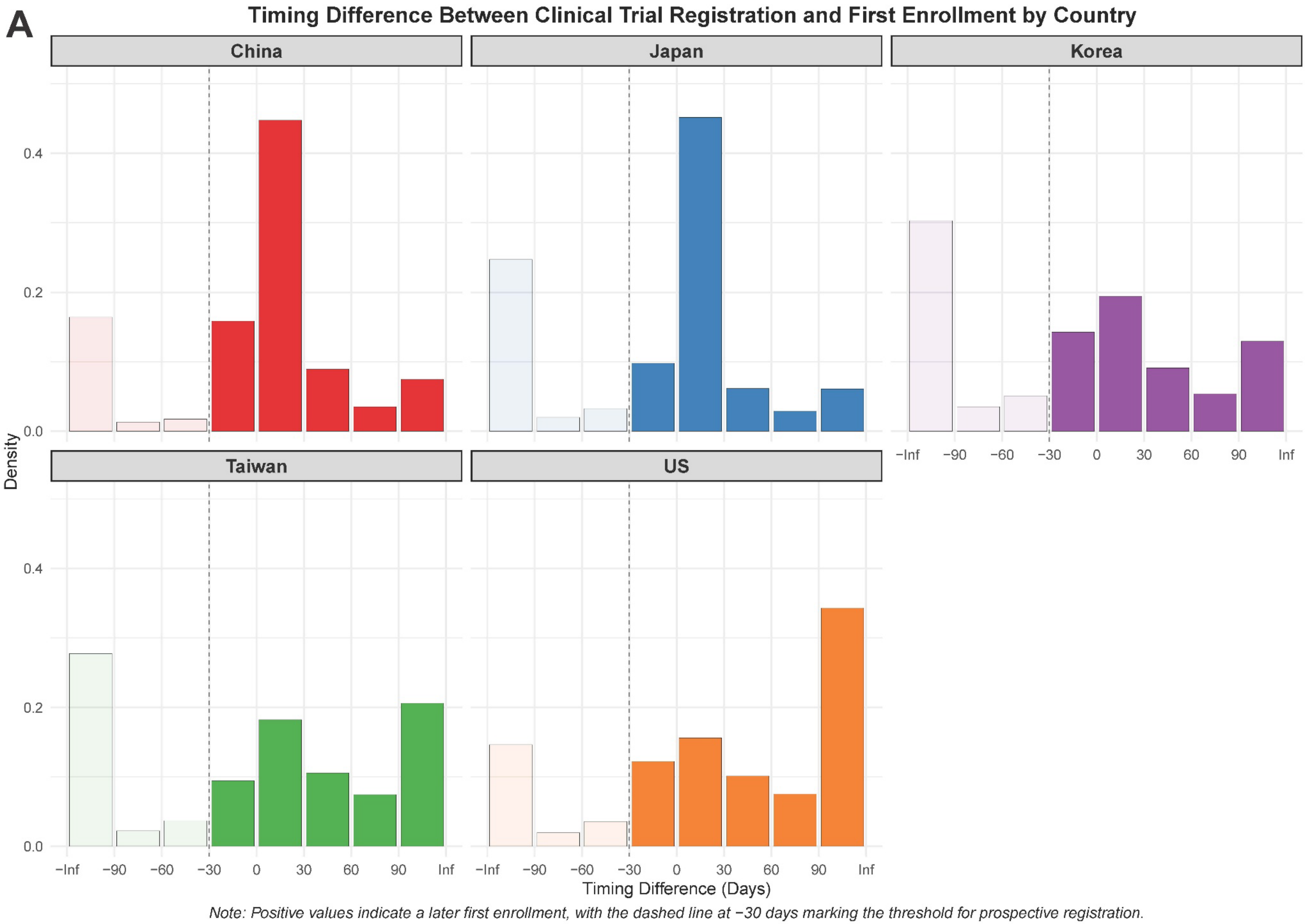


**Supplementary Figure 6.** (A) Distribution of time differences between trial registration and first participant enrollment for all clinical trials, integrating data over the last decade (2014–2023).

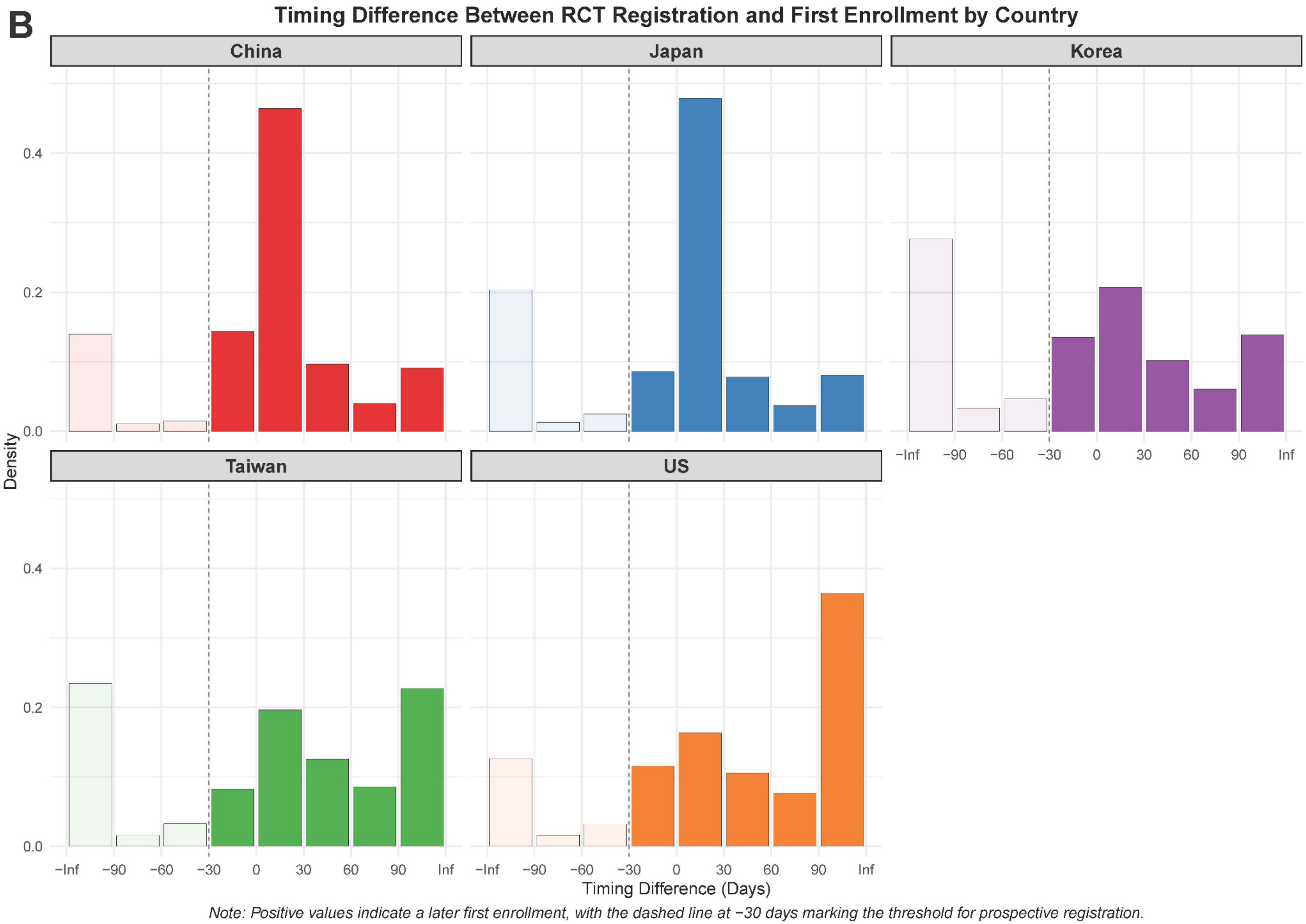


**Supplementary Figure 6.** (B) Distribution of time differences between trial registration and first participant enrollment for all RCTs, integrating data over the last decade (2014–2023). RCT, Randomized Controlled Trial.
